# Vaccine hesitancy and non-vaccination in an Irish paediatric outpatient population

**DOI:** 10.1101/2020.10.26.20219964

**Authors:** SO Whelan, F Moriarty, L Lawlor, K Gorman, J Beamish

**Affiliations:** Department of General Paediatrics, CHI at Temple Street, Dublin, Ireland; School of Pharmacy and Biomolecular Sciences, Royal College of Surgeons in Ireland, Dublin, Ireland; Department of Neurology and Clinical Neurophysiology, CHI at Temple Street, Dublin, Ireland; School of Medicine and Medical Sciences, University College Dublin, Dublin, Ireland

**Author notes:** Corresponding author –. Department of Paediatrics, Cork University Hospital, Cork, Ireland. **Contributorship Statement:** SOW conceived the study. SOW and JB designed the study. SOW and LL collected the data. FM conducted the analysis. All authors interpreted the findings. SOW, FM and KG drafted the manuscript. All authors critically revised and approved the final manuscript. JB oversaw the study process.

**Keywords:** vaccine hesitancy, vaccine refusal, childhood vaccinations, vaccine promotion, public health, immunisation

## Abstract

**Objective:** To administer the Parent Attitudes Childhood Vaccines (PACV) questionnaire to assess vaccine hesitancy and its relationship with non-vaccination.

**Design:** A cross-sectional study using the 15-item PACV questionnaire, with sociodemographic questions.

**Setting:** Outpatient department in a tertiary paediatric hospital, Dublin, Ireland.

**Participants:** Parents/caregivers of children attending general paediatric clinics.

**Main outcome measures:** PACV score and reported non-vaccination. We assessed sociodemographic factors associated with PACV score and accuracy of the PACV in predicting non-vaccination.

**Results:** In total, 436 participants completed the questionnaire. 5.5% of our population reported non-vaccination. HPV and MMR vaccines were the most commonly cited vaccines of concern (11.5% and 6.7% respectively) and autism spectrum disorder was the most commonly side effect of concern (4.3%). Mean PACV score was 26.9 (SD 19.1), with a significant difference between non-vaccinators and vaccinators (53.2 vs 25.3, p<0.001). Safety and efficacy concerns were the major contributor to non-vaccination. 14.4% of our population were vaccine-hesitant using the conventional cut-off score, which increased to 22% when using an optimal cut-off which maximised sensitivity and specificity. The accuracy of the PACV score to identify non-vaccination was good (area under the ROC curve = 0.827) and the optimal cut-off had a high negative predictive value (98.5%).

**Conclusions:** PACV identified non-vaccination with high accuracy in our population. It may be useful to screen vaccine hesitant parents who could benefit from interventions to improve uptake.

## Introduction

Vaccination is amongst the most successful public health interventions, second only to the provision of clean drinking water in reducing the burden of infectious diseases [1]. Globally, child mortality from vaccine-preventable diseases has fallen by more than two-thirds in the last three decades, in large part due to the increased availability of vaccinations [2]. While this benefit is preferentially seen in the developing world, it is estimated that childhood vaccination of a single year birth cohort in the United States of America (USA) prevented 42,000 deaths, and 20 million disease cases [3]. This does not include the deaths from Human Papilloma Virus (HPV)-related cancers which will ultimately result from the introduction of HPV vaccination [4].

Despite these successes, public confidence in vaccination has fallen worldwide, with associated reductions in vaccination rates in the United Kingdom (UK) and Ireland [5,6], resulting in disease outbreaks [7,8]. Even modest falls in vaccine uptake have significant public health and economic consequences, with one model suggesting a 5% reduction in measles, mumps rubella (MMR) uptake is associated with a 3-fold annual increase in cases [9]. This drop in uptake is multifactorial, but chief among the causes is vaccine hesitancy, recognised by the World Health Organisation (WHO) in 2019 as one of biggest threats to global health [10].

There are variations in how vaccine hesitancy is defined. The WHO’s Strategic Advisory Group of Experts on Immunization (SAGE) Working Group defines it as ‘*as a delay in acceptance, or refusal of vaccines, despite their availability*’ [11]. Other groups have created alternative models of vaccine behaviour, several of which divide the population into categories depending on attitudes and responses to vaccination. The common thread through these theories is the concept that a population’s attitudes towards vaccination exists on a continuum between demand for all vaccines on the one hand, and rejection of all on the other. The cohort between the extremes are ‘vaccine hesitant’. Determinants of vaccine hesitancy have been summarised using the WHO’s 3 Cs model – confidence in the vaccines and the system that administers them, complacency regarding the risks of vaccine preventable diseases, and convenience of physically getting the vaccination These are all subject to contextual, individual or vaccine specific influences [11].

This is a complex and context specific group, with a wide variety of views which are time, geography and vaccine dependent. Parents may choose to accept some childhood vaccines completely, delay or decline doses of some, and decline others outright. They may make different choices for different doses and different children, as their view changes over time. Local factors, such as outbreaks of vaccine preventable diseases may also encourage vaccination, while locally reported vaccine side effect fears may have the opposite effect. In addition, vaccine hesitancy cannot be solely based on vaccine uptake, as it is possible for vaccines to be accepted, while still holding concerns regarding this choice. Given this heterogeneity, it is perhaps unsurprising that the label of vaccine hesitancy is variably applied in different studies, impacting the comparability of results between studies [12,13].

For paediatricians and all healthcare professionals, understanding, targeting and influencing vaccine-hesitant cohorts towards positive vaccination choices is an important but challenging role. While consistently rated as one of the most trusted sources of information on vaccines [14], physicians frequently report feeling unprepared for dealing with vaccine hesitancy.

While clearly a population who need attention from healthcare professionals, identifying vaccine-hesitant parents is challenging. A recent systematic review noted there have been no experimental studies of interventions to enhance vaccine uptake in young children, which may be partially due to difficulty in identifying participants with hesitant views [15]. In 2011, Opel and colleagues developed the Parent Attitudes about Childhood Vaccines (PACV) survey, as a tool to assess parental vaccine hesitancy [16]. This has been validated to identify vaccine hesitance, and predict non-vaccination [17,18]. The PACV has since been adapted for use in other populations [19–21], translated into multiple languages [22,23] and used as an outcome in experimental studies [24,25].

Vaccine hesitancy is context-specific, varying across regions and healthcare systems, however the PACV has yet to be evaluated in a Western European setting. Our aim was to administer the PACV to assess vaccine hesitancy and its relationship with non-vaccination among parents and caregivers of children attending outpatient clinics in an Irish paediatric hospital. We sought to quantify the rates of vaccine hesitancy and non-vaccination, to investigate vaccination-related concerns, and to assess and optimise the performance of the PACV to predict non-vaccination in our population.

## Methods

### Study design, setting, and participants

This is a cross-sectional study, reported in line with the Strengthening Reporting of Observational Studies in Epidemiology (STROBE) statement. It was conducted from September to December 2018 in the general paediatric outpatients department of Children’s Health Ireland at Temple Street, Dublin, Ireland, a national acute hospital with providing secondary and tertiary care to children. General paediatric referrals are drawn from the north Dublin catchment area, with referrals largely drawn from general practitioners, subspecialist paediatricians and paediatric surgeons. Parents and caregivers accompanying a child to a clinic visit were eligible for participation in the study. All parents and caregivers were approached when checking in for their clinic visit and invited to read a participant information leaflet about the study and to complete the questionnaire, if interested. One response per child was invited, so if a child was accompanied by two adults, only one was invited to participate. The questionnaire was completed in the waiting room without assistance unless requested. Exclusion criteria included (i) parent/caregiver under 18 years of age, (ii) attending clinic with an interpreter, or (iii) had previously participated. Completed questionnaires were returned to a closed box, and were not seen by the attending clinician. This study was approved by the Children’s Health Ireland at Temple Street Ethics Research Committee (Reference: 18.071).

### Variables

The study tool (included in the supplementary materials) was a questionnaire consisting of the 15-item PACV questionnaire, demographic questions, vaccination status/intention for the participant’s oldest child, and white-space questions about vaccine-specific and side-effect concerns. The draft questionnaire was adapted for an Irish population following a pilot among a convenience sample of five parents/caregivers who met the inclusion criteria. Sociodemographic questions about income level and marital status were removed as these were felt to be overly intrusive by our pilot group and would likely have impacted on levels of participation. The word ‘shot’ was replaced with ‘vaccine’ throughout to match Irish vernacular.

The PACV is a 15-item questionnaire, divided into 3 domains, behaviour (2 items), safety and efficacy (4 items) and general attitudes and trust (9 items). There are three response designs in the PACV; dichotomous, a 5-point Likert scale, and an 11-point scale (e.g. ranging from ‘0 – Not sure at all’ to ‘10 – completely sure’’). All items are assigned a numeric score. A simple linear transformation was used to convert raw score to a 0-100 scale, with higher scores indicating increased hesitance. Participants were dichotomised as ‘non-hesitant’ with a score <50, and ‘hesitant’ if score was ≥50, in line with previous research [16] Scores were also calculated for each domain using a similar linear transformation.

The vaccination status of the participant’s oldest child was collected by self-report (with response options being received all vaccines, will receive all vaccines, will/has not received any vaccines or will/has not received certain vaccines). Participants were invited to disclose specific vaccines and specific side effects they had concern about. Participants’ age, level of educational attainment, and relationship to the child attending, and age of the participant’s oldest child were also collected. Education was defined as primary (i.e. up to approximately 8 years), partial or completed secondary (between 9 and 14 years), and tertiary (greater than 14 years). Relationship to child was defined as mother, father, grandparent, sibling or other.

### Statistical analysis

Statistical analysis was performed using Stata v14.0.with significance set at P-value < 0.05. Parametric tests were performed where the data were normally distributed, and non-parametric tests were employed if the data was not normally distributed. We summarised individual PACV item responses, domain scores, and total PACV score, and compared total score between vaccinators and non-vaccinators using a t-test with unequal variance. Univariate and multivariate linear regression were used to assess the association between participant characteristics (age of the participant, their relationship to the child, their educational attainment, age of their oldest child and whether vaccine/side effect concerns were report) and PACV score, estimating beta-coefficients with 95% confidence intervals (CIs). The ability of the PACV to predict non-vaccination was assessed. The area under the Receiver Operator Characteristic (ROC) curve was calculated, representing the accuracy with which the PACV score distinguished between non-vaccinators and vaccinators. This ranges from 0.5 (no better than chance) to 1.0 (perfect accuracy). We identified an optimal cut-off point for classifying hesitancy using three approaches: 1) the point nearest the top left corner of the ROC curve (indicating perfect sensitivity and specificity), 2) the point that maximises the product of the sensitivity and specificity, and 3) the point that maximises the sum of the sensitivity and specificity. Measures of diagnostic accuracy (sensitivity, specificity, positive and negative predictive values, and likelihood ratios for positive and negative results) with 95% CIs were calculated using the conventional cut-off of ≥50 for the PACV, and the optimal cut-off point identified.

## Results

This study included 436 participants (**Table 1**), with a mean age of 38.1 (SD 7.5) years, the majority of whom were parents (97.3%) and had completed third-level education (65.9%). Twenty-two participants (5.5%) reported their oldest child either had or would not receive some or all vaccines. A childhood vaccine that caused concern was reported by 21.6%, most frequently HPV and MMR (11.5% and 6.7%, respectively). Concerns were reported more frequently among non-vaccinators (72.7% versus 19.9% among vaccinators). A total of 16.1% parents/caregivers reported a potential vaccine side effect that they were concerned about, the most common being autism spectrum disorder (ASD) (4.3%), allergy/anaphylaxis or an unspecified reaction (2.3%) (**Supplementary Figures 1 and 2**).

**Table 1.** Descriptive characteristics of participants

| Characteristic | Total<br>(n=436) | Vaccinated<br>(n=391) | Non-vaccinated<br>(n=22) |
| --- | --- | --- | --- |
| **Age of participant (years), Mean (SD)** | 38.1 (7.53) | 38.4 (7.38) | 37.0 (8.80) |
| **Relationship, n (%)** |  |  |  |
| Mother | 344 (79.1) | 307 (78.5) | 19 (86.4) |
| Father | 79 (18.2) | 73 (18.7) | 3 (13.6) |
| Other | 11 (1.4) | 11 (2.8) | 0 (0.0) |
| **Educational attainment, n (%)** |  |  |  |
| Primary | 20 (4.7) | 19 (4.9) | 0 (0.0) |
| Some secondary | 34 (8.0) | 30 (7.8) | 2 (9.5) |
| All secondary | 91 (21.4) | 82 (21.2) | 4 (19.0) |
| Tertiary | 280 (65.9) | 255 (66.1) | 15 (71.4) |
| **Age of oldest child (years), Mean (SD)** | 9.9 (7.01) | 10.0 (6.94) | 9.9 (7.32) |
| **Concerned by at least one childhood vaccine, n (%)** | 94 (21.6) | 78 (19.9) | 16 (72.7) |
| **Reported a side effect of concern, n (%)** | 59 (13.5) | 44 (11.3) | 13 (59.1) |
| **Reported vaccination status of oldest child, n (%)** |  |  |  |
| Received all vaccinations | 348 (84.3) | 348 (89.0) |  |
| Intend to fully vaccinate | 43 (10.4) | 43 (11.0) |  |
| Not received any vaccines | 3 (0.7) |  | 3 (13.4) |
| Not receiving certain vaccines | 19 (4.6) |  | 19 (86.6) |

The mean PACV score was 26.9 (SD 19.1) and the distribution of scores is shown in **Figure 1**. The mean score for the Safety and Efficacy domain was 51.7 (SD 31.4), which was significantly higher than for the General Attitudes and Trust domain (20.0, SD 19.0), which was significantly higher than the Behaviour (8.7, SD 24.7) domains (p<0.001 for both, see **Supplementary Figure 3**). Vaccinators had a significantly lower mean PACV score compared to non-vaccinators (25.3 vs 53.2, p<0.001). The scoring of individual questions is shown in **Figure 2**. The question “*How concerned are you that your child might get a serious side effect from a vaccine?*” had the highest proportion of hesitant responses at 51.8%, while “*All things considered, how much do you trust your child’s doctor?*” had the lowest proportion of hesitant responses at 3.2%.

**Figure 1.**
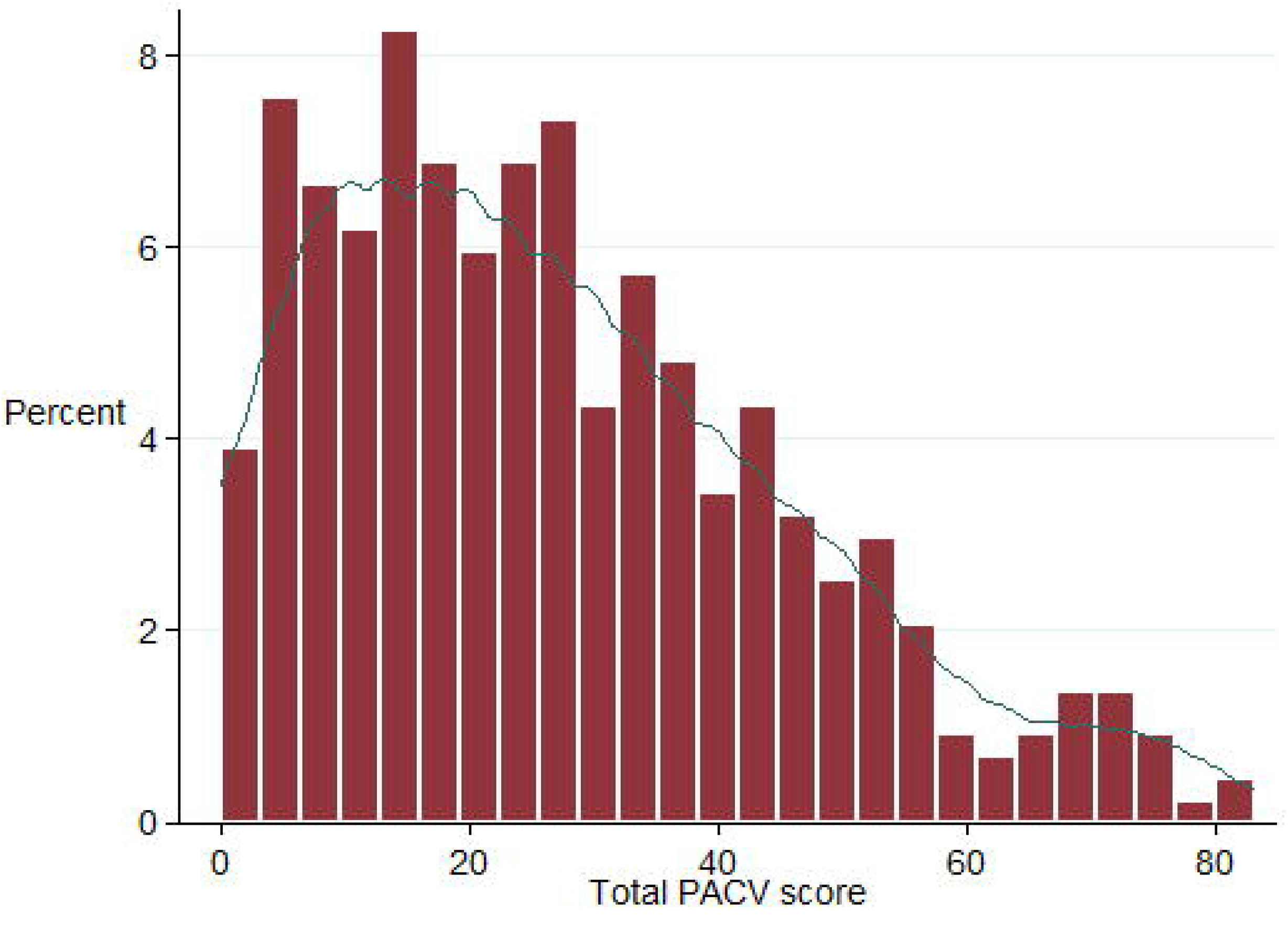
Distribution of PACV scores.

**Figure 2.**
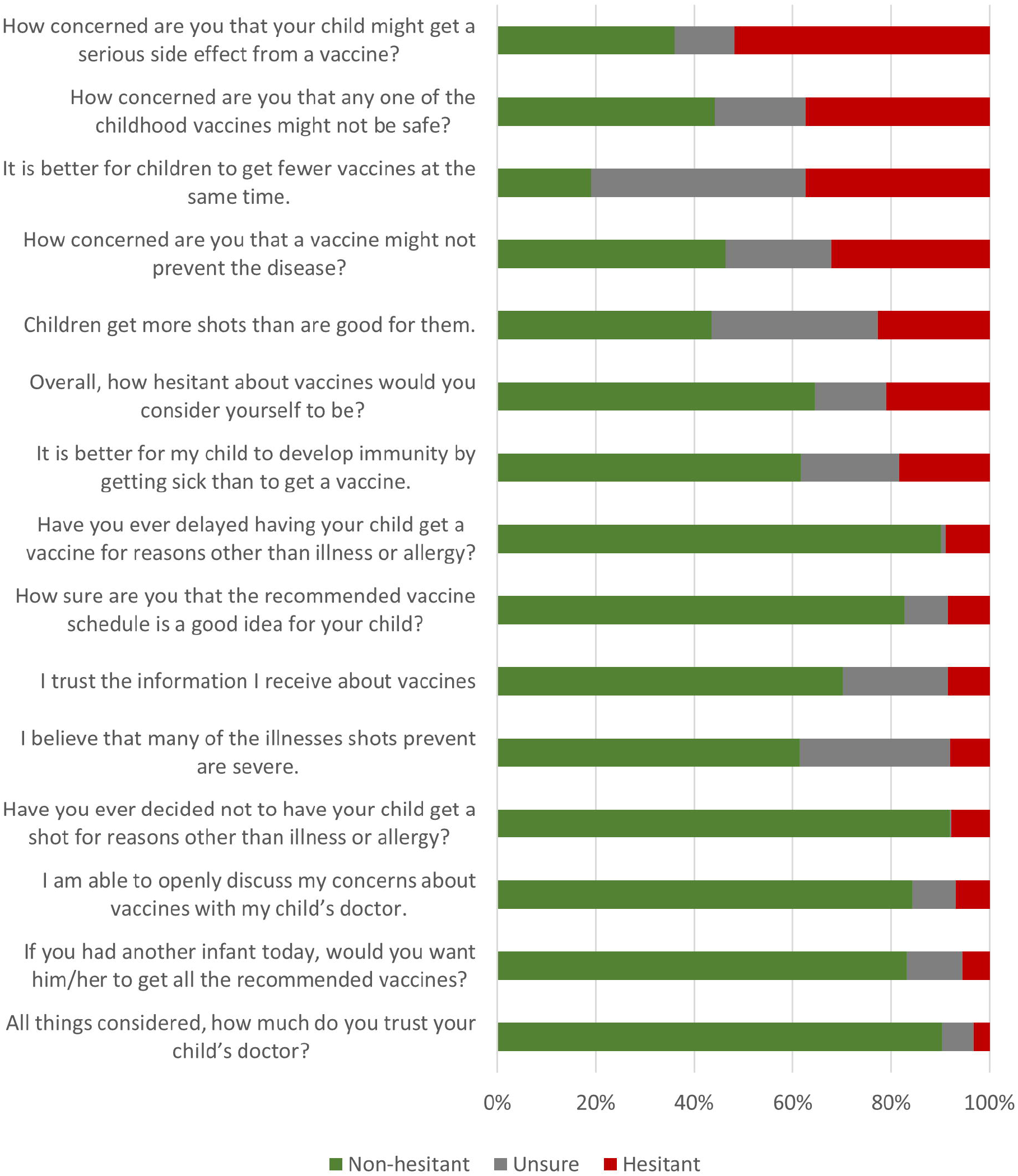
Distribution of responses for each of the 15 items within the PACV.

Examining factors associated with PACV score (**Table 2**), there was a statistically significant relationship between participant age and lower score, with each year increase in age corresponding to a decrease of −0.75 (95% CI −1.12 to −0.38) in PACV score, independent of other factors. Conversely, each year increase in the age of a participant’s oldest child was associated with an increase of 0.69 (95% CI 0.31 to 1.07). Reporting a childhood vaccine of concern was associated with an increase of 10.59 (95% CI 5.02 to 16.17), while reporting a side effect of concern also had a statistically significant relationship with higher score (8.24, 95% CI 1.49 to 14.99).

**Table 2.** Association of participant factors with PACV score from univariate (unadjusted) and multivariate (adjusted) linear regression

|  | Unadjusted |  | Adjusted |  |
| --- | --- | --- | --- | --- |
|  | Beta coefficient<br>(95% CI) | p value | Beta coefficient<br>(95% CI) | p value |
| **Age of participant, years** | -0.16 (-0.42, 0.09) | 0.207 | -0.75 (-1.12, -0.38) | <0.001 |
| **Relationship** |  |  |  |  |
| Mother (reference group) | 0.00 |  | 0.00 |  |
| Father | -3.51 (-8.19, 1.18) | 0.142 | 1.45 (-4.26, 7.16) | 0.618 |
| Other | 4.23 (-6.79, 15.26) | 0.451 | 9.04 (-10.52, 28.60) | 0.364 |
| **Educational attainment** |  |  |  |  |
| Primary | 5.29 (-3.50, 14.07) | 0.238 | 6.42 (-4.87, 17.72) | 0.264 |
| Some secondary | 1.36 (-5.53, 8.26) | 0.697 | 5.07 (-4.71, 14.86) | 0.308 |
| All secondary | 0.47 (-4.11, 5.05) | 0.841 | -0.06 (-5.85, 5.74) | 0.984 |
| Third level (reference group) | 0.00 |  | 0.00 |  |
| **Age of oldest child, years** | 0.32 (0.02, 0.63) | 0.039 | 0.69 (0.31, 1.07) | <0.001 |
| **Concerned by at least one childhood vaccine** | 11.29 (7.04, 15.53) | <0.001 | 10.59 (5.02, 16.17) | <0.001 |
| **Reported a side-effect of concern** | 11.98 (6.84, 17.12) | <0.001 | 8.24 (1.49, 14.99) | 0.017 |

The accuracy of the PACV score to identify non-vaccinators was good, with an area under the ROC curve of 0.827 (**Figure 3**). The optimal cut-off point was identified 41.67, which was the point closest to the top left corner of the ROC curve and also maximised both the product and sum of the sensitivity and specificity. This classified 96 (22%) of participants as hesitant, as opposed to 63 (14.4%) identified using the conventional cut-off point of ≥50, and measures of diagnostic accuracy for both are shown in **Supplementary Table 1**. The optimal cut-off point of 41.67 had a sensitivity of 77.3% (95% CI 54.6% to 92.2%) and a specificity of 81.3% (95% CI 77.1% to 85.1%). The positive predictive value of being classed as hesitant was 18.9% (95% CI 11.4% to 28.5%) and the negative predictive value of being classed as non-hesitant was 98.5% (95% CI 96.4% to 99.5%). The likelihood ratio for being classed as hesitant was 4.1 (95% CI 3.1 to 5.6) and for being classed as non-hesitant was 0.3 (95% CI 0.1 to 0.6).

**Figure 3.**
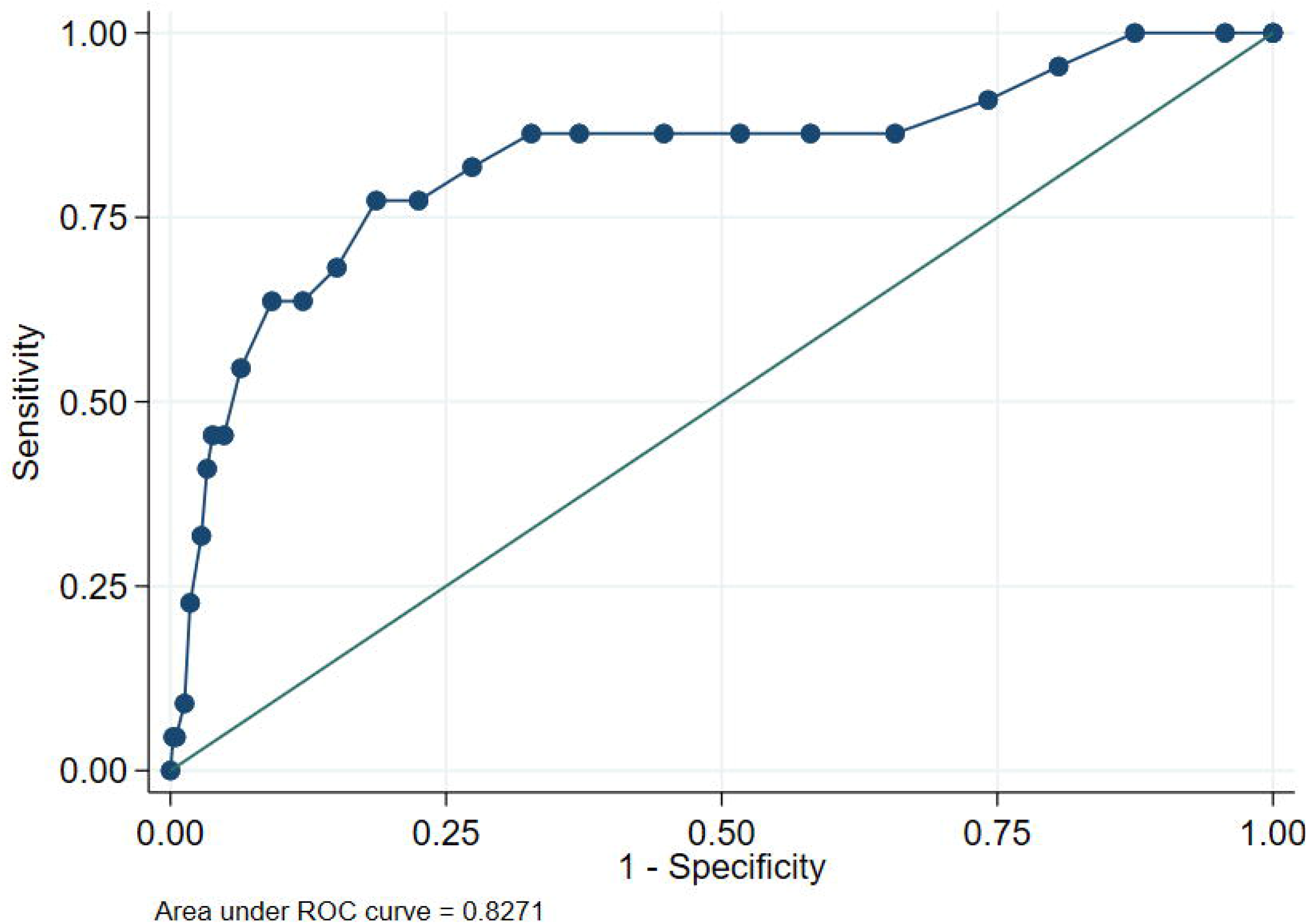
Receiver Operating Characteristic (ROC) curve for the PACV score in predicting non-vaccination.

## Discussion

Our study, one of the largest PACV studies to date, and the first in a Western European population, studying the accuracy of PACV in predicting vaccine hesitancy in a population of parents/caregivers attending a general paediatric outpatient. Employing/implementing a PACV score > 41 is associated with non-vaccination, with higher sensitivity and specificity that using a PACV score >50.

Nearly 15% of our population was vaccine-hesitant using the conventional PACV cut-off, similar to other populations including Quebec, Canada, (15%) and Indonesia (15.9%) [26,27]. The prevalence identified in our study is higher than in Iraq (9.9%) and the United Arab Emirates (12%) [23,28] and lower than Italy (34.7%) and several USA studies (>20%) [17,21,29]. This places Ireland in the middle of vaccine hesitancy estimates, which concurs with the 2018 Wellcome Global Monitor Report [30]. This had asked a sample of Irish people whether vaccines are important to have, and 91% had somewhat or strongly agreed.

We found that older participants had a lower PACV (a reduction of −0.75 (95%CI −1.12, −0.38 in PACV score associated with each year increase in age) which has been observed previously [22]. It is has been suggested that the higher levels of hesitance in younger parents may be due to increased use and the influence of social media in younger cohorts, which is predominantly anti-vaccination in outlook [15]. We also found that those with younger children were more likely to be vaccine-hesitant, contrasting with the Malaysian study which suggested first-time mothers were more vaccine-hesitant [22].

Our hesitancy rate was three times higher than the non-vaccination rate, suggesting there may be a significant proportion of parents who are choosing to vaccinate, but still have significant vaccine concerns. The national trends in vaccination rates in Ireland align with this finding. Human papilloma virus vaccine uptake was 85.5% in the 2011/2012 school year, but fell to 51% in 2016/2017, coinciding with a rise in anti-HPV vaccine publicity [31]. This experience has been mirrored internationally [32]. A concerted, multi-modal cross-sectoral alliance was built by Ireland’s National Immunisation Office to reverse this trend, resulting in an increase in the vaccination rates to 64.1% in 2017/2018 [33]. This experience illustrates how hesitant parents may be persuaded for or against vaccination, depending on the information they are provided with. More than one in ten participants in this study were still concerned about HPV vaccination today, emphasising the need for continued vigilance against future declines in vaccination.

As in the anti-HPV vaccine campaign, our results showed that safety and efficacy is the primary motivator for vaccine hesitance, with this domain having the highest score, with a mean of 51.7(31.4). This result mirrors other PACV research worldwide [20,27], and safety concerns are the most commonly cited barrier to vaccination [34]. Keeping children healthy is one of the most important considerations for parents, and choices can be based on a complex balancing of risks and benefits [35]. The fraudulent association between vaccines and ASD has been thoroughly debunked, however this side effect was the single most commonly cited in our population (4.3%). Similar to HPV, MMR vaccination rates declined sharply following this controversy in the late 1990s. In the intervening period, while immunisation levels have recovered, parental reservations regarding ASD have remained relatively unchanged [36]. Allaying these concerns is important for ensuring continued improvements in MMR uptake.

Over 90% of our participants trusted their doctor. Trust in healthcare professionals, and paediatricians in particular, has been consistently described as critical for vaccine uptake [14]. Indeed, some American surveys of parents of vaccination aged children suggest that the influence of the paediatrician is the single most powerful factor in vaccine acceptance [37]. This places professionals in a unique position to promote vaccination amongst their patient’s parents. Increasing numbers of paediatricians are caring for the children of parents who have refused a vaccine [38], but up to half of these may be convinced to accept vaccination with continued engagement [39]. It has become the recommendation or professional associations including the American Academy of Paediatrics to continue to engage with vaccine-hesitant parents, in the hopes of a positive vaccination decision in the future [40].

These consultations can be challenging. The provision of information about the safety of vaccines and the dangers of vaccine preventable diseases, while important, may not alone be sufficient to reduce hesitancy, change minds and increase vaccine uptake [41]. Communication is based on a mutual understanding that both parents and healthcare professionals are acting in what they consider to be the best interests of the child [42] It is crucial to engage with specific concerns raised by parents about specific vaccines for their child, and recognise that having concerns is a natural, even healthy response [43]. Evidence-based methods for vaccine promotion amongst parents include using a presumptive, rather than a participatory, approach to vaccine discussion, to make vaccine acceptance rather than refusal the default position [44]. For those who resist vaccination, motivational interviewing techniques have been shown to be effective, as has persistence with positive messaging over the course of the clinical relationship [44].

In addition, it has become increasingly apparent that the use of personal stories to illustrate the importance of vaccination and the dangers of vaccine-preventable diseases is an effective tool [45]. While anecdotes have traditionally been frowned upon by the medical community, they have played a key role in combatting the proliferation of first-hand accounts of perceived harms from vaccination, amplified by social media, that result in vaccine refusal. Empirical evidence has shown that anti-vaccination narratives can override the effect of statistical information and influence risk perception about vaccines, particularly when the narrative is highly emotional [46]. Similar strategies to combat these sentiments have been employed. For example, stories involving vaccine preventable disease experiences, uneventful vaccinations, or ‘conversions’ of previously anti-vaccination voices have all been used to effect in campaigns internationally [45]. Directing parents to pro-vaccination parent curated social media platforms may counter the anti-vaccination equivalents.

Applying these techniques in clinical practice can be time-consuming and daunting, which can have negative outcomes for both parents and healthcare professionals [47]. As such, targeting the interventions to where they are most needed is critical. Our results suggest the PACV is a robust, accurate tool to identify parents who are vaccine hesitant, as a focus for targeted vaccine promotion. We identified a novel cut-off point for hesitance to optimise the tool’s performance in detecting potential non-vaccinators. Ours is lower than the conventional cut-off point (41.7 versus 50), identifying 22% rather than 14.4% of our population as hesitant, optimising sensitivity and specificity. Using the PACV with our new cut-off point to target non-vaccinators, rather than an untargeted approach, we increase the likelihood of targeting a non-vaccinator from 1 in 20, to 1 in 5, assuming a non-vaccination rate of 5%, as in our population. In clinical practice, the PACV offers the realistic prospect of engaging parents who are at high risk of non-vaccination, in a timely fashion. Based on the negative predictive value, those identified as non-hesitant have a high probability (98.5%) of being vaccinators and therefore a clinician is unlikely to miss a non-vaccinator. In more time-constrained settings, the higher conventional cut-off point of 50 could be used. This would increase the specificity, classifying fewer people as hesitant with a higher proportion of these being non-vaccinators, but at the expense of lower sensitivity, resulting in more non-vaccinators being classified as non-hesitant.

Our study was limited by lack of access to vaccination data and relied on the use of self-reported vaccination, as have many previous vaccine hesitancy studies [4,22,23,27]. Reliability of self-report has only been evaluated in adults and for specified vaccinations [48,49], However parental report of adherence to childhood vaccination in general may be expected to be more accurate, especially for deliberate partial or complete non-vaccination. In the absence of national life-long registries however, self-report remains the practical option for vaccination information. We used vaccination status of the oldest child in our study, with a mean age of 9.9. This is older than many other PACV studies, and may affect recall. However with the HPV vaccine as a further vaccination choice that parents of these older children will still have to make, evaluating vaccine hesitancy in this group is vital. In fact, hesitancy in this group may be particularly important, given the anti-vaccination movement has focused largely on HPV vaccines. Secondly, our population were attending a paediatric clinic, and therefore may not be representative of a general population of children. For instance, there is evidence that children with chronic conditions, more prevalent in a hospital attending population than the community at large, may be under-vaccinated when compared with their peers [50]. Our results should not be assumed to apply to children attending other settings, including primary care or routine childhood screening for development or growth. However the findings are informative for clinicians who are encountering a population similar to that in this study. Our non-vaccination rates are lower than national and local averages [6,33], which may reflect this as well as social desirability bias. The representativeness of our population (e.g. 79.1% being mothers) may limit the generalisability of our results to others involved in vaccination decisions. That said, there are indications that mothers are the predominant decision makers in terms of their children’s health [51]

## Conclusion

In conclusion, we found that vaccine hesitancy was relatively common in our population, and that the PACV identified non-vaccination with high accuracy. Using a lower cut-off point of >41.7 may optimise use of the PACV compared to the conventionally used cut-off point of 50. Therefore, the PACV may be a useful tool for paediatricians to effectively screen their clinic for vaccine hesitant-parents and to support vaccination uptake over the course of childhood.

## Data Availability

Data available as per manuscript only.

