## Supplementary Files for "Vaccine hesitancy and non-vaccination in an Irish paediatric outpatient population"

Supplementary File 1 – Questionnaire

**PERCEPTIONS ABOUT CHILDHOOD VACCINES SURVEY**

1. **Have you ever delayed having your child get a vaccine for reasons other than illness or allergy? [Please tick your response in the space provided]**

Yes ____	No ____	Don’t know ____

2. **Have you ever decided not to have your child get a shot for reasons other than illness or allergy? [Please tick your response in the space provided]**

Yes ____	No ____	Don’t know ____

3. **How sure are you that the recommended vaccine schedule is a good idea for your child?**

0	1	2	3	4	5	6	7	8	9	10

0 = Not at all sure	10 = Completely sure

4. **If you had another infant today, would you want him/her to get all the recommended vaccines? [Please tick your response in the space provided]**

Yes ____	No ____	Don’t know ____

5. **Overall, how hesitant about vaccines would you consider yourself to be? [Please tick your response in the space provided]**

Not at all hesitant ____

Not that hesitant ____

Not sure ____

Somewhat hesitant ____

Very hesitant ____

6. **Children get more shots than are good for them. [Please tick your response in the space provided]**

Strongly agree ____	Agree ____	Not sure ____	Disagree ____	Strongly disagree ____

7. **I believe that many of the illnesses shots prevent are severe. [Please tick your response in the space provided]**

Strongly agree ____	Agree ____	Not sure ____	Disagree ____	Strongly disagree ____

9. **It is better for my child to develop immunity by getting sick than to get a vaccine. [Please tick your response in the space provided]**

Strongly agree ____	Agree ____	Not sure ____	Disagree ____	Strongly disagree ____

10. **It is better for children to get fewer vaccines at the same time. [Please tick your response in the space provided]**

Strongly agree ____	Agree ____	Not sure ____	Disagree ____	Strongly disagree ____

11. **How concerned are you that your child might get a serious side effect from a vaccine? [Please tick your response in the space provided]**

Not at all concerned ____

Not that concerned ____

Not sure ____

Somewhat concerned ____

Very concerned ____

12. **How concerned are you that any one of the childhood vaccines might not be safe? [Please tick your response in the space provided]**

Not at all concerned ____

Not that concerned ____

Not sure ____

Somewhat concerned ____

Very concerned ____

13. **How concerned are you that a vaccine might not prevent the disease? [Please tick your response in the space provided]**

Not at all concerned ____

Not that concerned ____

Not sure ____

Somewhat concerned ____

Very concerned ____

14. **I trust the information I receive about vaccines. [Please tick your response in the space provided]**

Strongly agree ____	Agree ____	Not sure ____	Disagree ____	Strongly disagree ____

15. **I am able to openly discuss my concerns about vaccines with my child’s doctor. [Please tick your response in the space provided]**

Strongly agree ____	Agree ____	Not sure ____	Disagree ____	Strongly disagree ____

16. **All things considered, how much do you trust your child’s doctor?**

0	1	2	3	4	5	6	7	8	9	10

0 = Do not trust at all	10 = Completely trust

**What age are you?** _____

**Which of the following best describes your relationship to the child you are accompanying today? [Please tick your response in the space provided]**

Mother ___

Father ____

Grandparent____

Sibling ___

Other ____ [Please describe ______________________________________]

**What is the highest level of education you have undertaken? [Please tick your response in the space provided]**

Primary School _____

Some secondary school _______

Leaving Certificate/Secondary school leaver’s exam _________

Third level (University/College, or similar) _______

Age of your oldest child: ___________

**Vaccination status of oldest child: [Please tick your response in the space provided]**

- My child has received all of their vaccinations
- I intend to fully vaccinate my child
- My child has not received, or will not receive, any vaccines
- I will not give my child certain vaccines

**Are there any vaccines you have concerns about your child receiving? If so, which ones, and why?**

______________________________________________________________________

______________________________________________________________________

________________________________________________________________________

________________________________________________________________________

**What side effects of vaccines are you concerned about?**

________________________________________________________________________

________________________________________________________________________

________________________________________________________________________

________________________________________________________________________

Percentage of participants

|  | Percent |
| --- | --- |
| HIB | 0.22936% |
| Men C | 0.22936% |
| TB | 0.22936% |
| Varicalla | 0.22936% |
| Men B | 0.45872% |
| BCG | 1.14679% |
| Rotavirus | 1.14679% |
| All vaccines | 1.83486% |
| MMR | 6.65138% |
| HPV | 11.4679% |

Supplementary figure 1. Frequency of specific childhood vaccines of concern reported by two or more participants

Percentage of participants

|  | Percent |
| --- | --- |
| Autoimmune disorder | 0.45872% |
| Death | 0.45872% |
| ADHD | 0.45872% |
| Rash | 0.45872% |
| Asthma | 0.68807% |
| Sleep disorder | 0.91743% |
| Learning disability | 0.91743% |
| Pain | 1.37615% |
| Fever | 1.37615% |
| Allergy/Anaphylaxis/Reaction | 2.29358% |
| Autism | 4.3578% |

Supplementary figure 2. Frequency of vaccine side-effects of concern reported by two or more participants

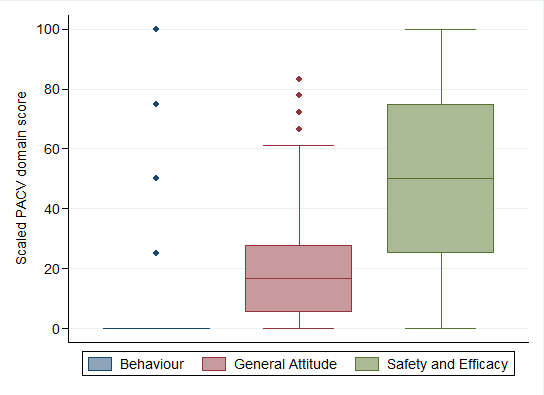

Supplementary figure 3. Box plot of the distribution of score across Behaviour, General Attitude, and Safety and Efficacy PACV domains.

Supplementary table 1. Measures of diagnostic accuracy for PACV to classify non-vaccination using optimal and conventional cut-off points

|  | **Estimate (95% confidence interval)** | |
| --- | --- | --- |
|  | **Optimal cut-off point (41.67)**  **(n=96)** | **Conventional cut-off point (50)**  **(n=63)** |
| **Sensitivity** | 77.3% (54.6%, 92.2%) | 63.6% (40.7%, 82.8%) |
| **Specificity** | 81.3% (77.1%, 85.1%) | 88.0% (84.3%, 91.0%) |
| **Positive predictive value** | 18.9% (11.4%, 28.5%) | 23.0% (13.2%, 35.5%) |
| **Negative predictive value** | 98.5% (96.4%, 99.5%) | 97.7% (95.6%, 99.0%) |
| **Likelihood ratio +** | 4.1 (3.1, 5.6) | 5.3 (3.5, 8.0) |
| **Likelihood ratio -** | 0.3 (0.1, 0.6) | 0.4 (0.2, 0.7) |
